# Increasing Numbers of Non-communicable Disease Co-morbidities: Major Risk Factors for Hospitalization among a Cohort of People with HIV and COVID-19 Coinfection

**DOI:** 10.1101/2021.08.03.21261560

**Authors:** Michael D. Virata, Sheela V. Shenoi, Joseph Ladines-Lim, Merceditas S. Villanueva, Lydia A. Barakat

**Author notes:** All authors contributed significantly to the study design and to the collection, organization, and analysis of the data, and participated in the preparation of this report. The authors have approved the final version for submission.

## Abstract

Data regarding coronavirus disease (COVID-19) infection complications among people with HIV (PWH) are expanding but sometimes conflicting. This report presents the results of a retrospective review of 103 patients at a single urban academic health center with confirmed severe acute respiratory syndrome coronavirus 2 (SARS-CoV-2) infection. The study showed that hospitalization is related to host factors such as age 65 years or older with an increasing number of specific non-communicable disease comorbidities, and not to HIV-attributable factors.

## Introduction

The first laboratory-confirmed case of coronavirus disease (COVID-19) secondary to severe acute respiratory syndrome coronavirus 2 (SARS-CoV-2) infection in the United States (US) came to the attention of the Centers for Disease Control and Prevention (CDC) on January 21, 2020. A mere 12 months later, cases had reached 24.5 million, with over 500,000 deaths recorded.[1]

Initial reports from Wuhan, China, identified a number of risk factors for severe COVID-19 and mortality. Among these factors were older age, male gender, and co-morbidities such as hypertension, diabetes, chronic cardiovascular and respiratory disease, and immunosuppressed conditions.[2] At that time, it was not known whether clinically stable persons with HIV (PWH) who were diagnosed with COVID-19 would be more likely to be hospitalized or to develop severe illness.

Several very early case series showed that clinical outcomes among PWH were not any worse than those for patients without HIV,[2,3,4] and found no excess risk of morbidity and mortality in symptomatic SARS-CoV-2 co-infected patients with fully suppressed HIV, compared with HIV-uninfected patients.[5,6] Similarly, a US study matched 21 HIV-infected against 42 HIV-uninfected patients, all hospitalized for COVID-19, and noted no significant difference in clinical course or outcomes.[7] A few months into the pandemic, there was more evidence from several areas in the US that HIV was not being identified as a risk factor for hospitalization.[8] However, a recent systematic review of 25 published studies of PWH co-infected with SARS-CoV-2 demonstrated that among 252 patients, 65% were hospitalized and 17% required admission to an intensive care unit (ICU).[9]

To gain a better understanding of the risk factors for hospitalization among PWH infected with SARS-CoV-2, a cohort study comparing ambulatory and hospitalized co-infected patients at a single urban academic center was made during the first year of the pandemic.

## Methods Used in the Study

### Study Design and Population

The study involved a retrospective chart review of PWH 18 years old or over who had a laboratory-confirmed SARS-CoV-2 infection between January 21, 2020, and January 20, 2021, using reverse transcription polymerase chain reaction (RT-PCR) test results. All patients received their primary care at two HIV ambulatory clinics within the Yale–New Haven Hospital (YNHH) academic medical center in New Haven, Connecticut. Eligible patients were divided into hospitalized and ambulatory groups.

### Data Sources and Analysis

A standardized form was used in abstracting data from electronic medical records (Epic Systems Corporation) including demographic information, co-morbidities, and clinical parameters such as body mass index (BMI), as well as HIV-specific information including AIDS diagnosis, antiretroviral therapy (ART), CD4 cell count and HIV viral load (HIVVL) before admission. Among the COVID-specific information items were COVID-19-related symptoms, reverse transcription polymerase chain reaction (RT-PCR) test results, and COVID-19 management protocols including specific SARS-CoV-2 therapies. Predetermined clinical outcomes included the need to escalate to ICU level of care, and all-cause mortality. HIVVL suppression was defined as <200 copies per milliliter (copies/ml) of blood; history of AIDS, as having had an opportunistic infection or a CD4 count of <200 cells/mm^3^ or both; and obesity, as BMI>30.

Continuous and categorical variables were summarized as medians or means, and percentages as appropriate, and bivariate analysis comparing hospitalized with nonhospitalized patients using SPSS Version 26.0 was performed to identify factors that differentiated between the two groups. Differences with a *p*-value less than 0.05 were significant. Multivariable backward logistic regression identified independent correlates for hospitalization.

The study was approved by the Yale University Institutional Review Board and the requirement for an informed consent was waived for this retrospective review. All patient records and pertinent information were de-identified prior to the analysis.

## Results

### Demographics

Among 1,469 PWH receiving care at the two clinics, 103 (7%) had confirmed COVID-19. The median age was 56 (interquartile range [IQR] 45–62) years—72 PWH (69.9%) were >50 years old, while 17.5% were >65 years old—and 46.6% were women, 43.7% were African Americans, and 16.5% were Latinx (**Table 1**). The median CD4 count was 735 cells/mm^3^ (IQR 434–928), 98.1% of the patients were prescribed ART, and 92.2% had HIVVL suppression. Among the 88% with co-morbidities, 51.5% had hypertension; 43.7%, obesity; 34%, underlying chronic lung disease; 27.2%, cardiovascular disease; 27.2%, diabetes mellitus; and 22.3%, chronic kidney disease. While 56.3% of PWH were former smokers, active use of tobacco was documented in 22.3%, alcohol in 27.2%, and illicit substances in 13.6%.

**Table 1.** Demographics and Clinical Characteristics of PWH with SARS-CoV-2, 1/21/2020-1/20/2021 (n=103)

|  | Total (n=103) | Ambulatory (n=69) | Hospitalized (n=34) | p-value | OR (95% CI) |  |
| --- | --- | --- | --- | --- | --- | --- |
| **Demographics** |  |  |  |  |  |  |
| Median age (IQR) | 56 (45–62) | 55 (42.5–59) | 58 (52.3–67.3) | 0.40<sup>#</sup> |  |  |
| Age≥50 yrs, n (%) | 72 (69.9) | 45 (65.2) | 27 (79.4) | 0.14 |  |  |
| Age>65 yrs, n (%) | 18 (17.5) | 8 (11.6) | 10 (29.4) | **0.025** | **3.18 (1.12–9.01)** |  |
| Women, n (%) | 48 (46.6) | 32 (46.4) | 16 (47.1) | 0.95 |  |  |
| African Americans, n (%) | 45 (43.7) | 30 (43.5) | 15 (44.1) | 0.95 |  |  |
| Latinx, n (%) | 17 (16.5) | 13 (18.8) | 4 (11.8) | 0.27 |  |  |
| **HIV History** |  |  |  |  |  |  |
| Median years living with HIV (IQR) | 16.5 (9–23.8) | 17 (8–25) | 16 (10–23) | 0.83<sup>#</sup> |  |  |
| History of AIDS, n (%) | 46 (44.7) | 30 (43.5) | 16 (47.1) | 0.75 |  |  |
| History of CD4<200 cells/mm<sup>3</sup>, n (%) | 43 (41.8) | 28 (40.5) | 15 (44.1) | 0.77 |  |  |
| History of OI, n (%) | 24 (23.3) | 18 (26.1) | 6 (17.6) | 0.25 |  |  |
| On ART, n (%) | 101 (98.1) | 67 (97.1) | 34 (100) | 0.32 |  |  |
| Median CD4 count (IQR) | 735 (434–928) | 718 (397–931) | 784 (466–924) | 0.67<sup>#</sup> |  |  |
| Proportion VL suppressed (VL<200), n (%) | 95 (92.2) | 65 (95.2) | 30 (88.2) | 0.25 |  |  |
| **Co-morbidities** |  |  |  |  |  |  |
| Active smoking, n (%) | 23 (22.3) | 15 (21.7) | 8 (23.5) | 0.84 |  |  |
| Former smoking, n (%) | 58 (56.3) | 37 (53.6) | 21 (61.8) | 0.43 |  |  |
| Active substance use, n (%) | 14 (13.6) | 6 (8.7) | 8 (23.5) | **0.039** | **3.2 (1.0–10.2)** |  |
| Active alcohol use, n (%) | 28 (27.2) | 19 (27.5) | 9 (26.5) | 0.91 |  |  |
| Diabetes mellitus, n (%) | 28 (27.2) | 14 (20.3) | 14 (41.2) | **0.025** | **2.75 (1.1–6.8)** |  |
| Median HgbA1C (IQR) (n=50) | 7.8 (6.4–8.5) | 7.1 (6.2–8.3) | 8.3 (6.6–9.0) | 0.20<sup>#</sup> |  |  |
| Chronic lung disease, n (%) | 35 (34) | 18 (26.1) | 17 (50) | **0.016** | **2.83 (1.20–6.7)** | **3.35 (1.28–8.72)** |
| Chronic kidney disease, n (%) | 23 (22.3) | 10 (14.5) | 13 (38.2) | **0.007** | **3.65 (1.39–9.57)** |  |
| Cardiovascular disease, n (%) | 28 (27.2) | 13 (18.8) | 15 (44.1) | **0.007** | **3.4 (1.37–8.42)** | **3.4 (1.27–9.12)** |
| Hypertension, n (%) | 53 (51.5) | 31 (44.9) | 22 (64.7) | 0.059 | 2.25 (0.96–5.25) |  |
| Hepatitis C, n (%) | 23 (22.3) | 11 (15.9) | 12 (35.3) | **0.027** | **2.88 (1.1–7.47)** |  |
| Median BMI (IQR) | 28.9 (25.2–34.9) | 29.3 (25.7–35.0) | 26.8 (24.7–36.0) | 0.30<sup>#</sup> |  |  |
| Obesity (BMI>30), n (%) | 45 (43.7) | 32 (46.4) | 13 (38.2) | 0.43 |  |  |
| Any co-morbidity, n (%) | 93 (90.3) | 62 (89.9) | 31 (91.2) | 0.83 |  |  |
| Number of co-morbidities | 3 (1–4) | 2 (1–3) | 4 (2–5) | **0.001\*** | **1.61 (1.22–2.13)** |  |

\*Logistic regression; <sup>#</sup>Mann Whitney U; PWH-persons with HIV

### Characterization of Hospitalized PWH

Thirty-four PWH (33.0%) were hospitalized and 69 (67.0%) were ambulatory. Hospitalized PWH represented <1% of all COVID-19-related admissions to YNHH during the study period. In the bivariate analysis, age and multiple co-morbidities were significant. In the adjusted analysis (**Table 1**), on the other hand, those who were hospitalized were more likely to be at least 65 years old (odds ratio [OR] 3.11, 95% confidence interval [CI] 0.97–9.98) and to have chronic lung disease (OR 3.35, 95% CI 1.28–8.72) or cardiovascular disease (OR 3.4, 95% CI 1.27–9.12). Moreover, incremental numbers of co-morbidities were associated with hospitalization (OR 1.61, 95% CI 1.22–2.13). AIDS history and last CD4 count were not associated with hospitalization. There was no significant difference in ART coverage or HIVVL suppression between inpatients and outpatients.

Among those who were hospitalized (**Table 2**), the most frequently prescribed inpatient therapies for COVID-19 were hydroxychloroquine (47.1%), tocilizumab (23.5%), and remdesivir and steroids (14.7%). A small proportion (14.7%) received lopinavir/ritonavir or atazanavir. During hospitalization, five patients (14.7%) required escalation to ICU level of care, six patients (17.6%) needed intermediate (step-down) care; and four (11.8%) had to be placed on mechanical ventilation. The median length of stay was nine days (IQR 3–15). Only one patient (2.9%) died within 30 days of hospital admission. In a subset of hospitalized patients (n=15), repeat CD4 counts revealed a decreased median of 417.8 (IQR 186–617) cells/mm^3^. Repeat HIVVL testing done for 59% of patients showed full viral suppression in 95% of that subset (**Table 2**).

**Table 2.**
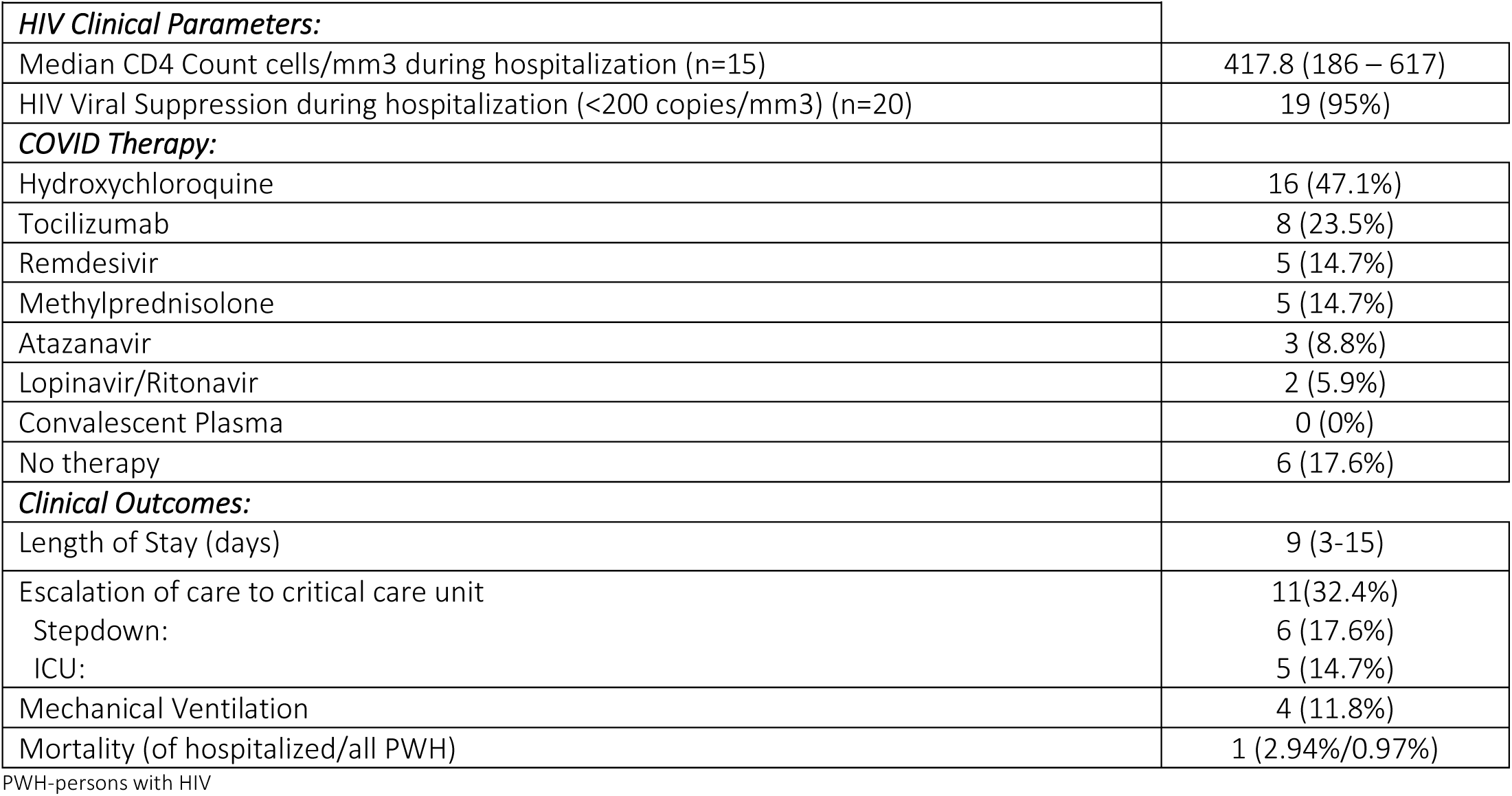
Characteristics of Hospitalized PWH (n=34)

| Table 2. Characteristics of Hospitalized PWH (n=34) |  |
| --- | --- |
| **HIV Clinical Parameters:** |  |
| Median CD4 Count cells/mm<sup>3</sup> during hospitalization (n=15) | 417.8 (186 – 617) |
| HIV Viral Suppression during hospitalization (<200 copies/mm<sup>3</sup>) (n=20) | 19 (95%) |
| **COVID Therapy:** |  |
| Hydroxychloroquine | 16 (47.1%) |
| Tocilizumab | 8 (23.5%) |
| Remdesivir | 5 (14.7%) |
| Methylprednisolone | 5 (14.7%) |
| Atazanavir | 3 (8.8%) |
| Lopinavir/Ritonavir | 2 (5.9%) |
| Convalescent Plasma | 0 (0%) |
| No therapy | 6 (17.6%) |
| **Clinical Outcomes:** |  |
| Length of Stay (days) | 9 (3-15) |
| Escalation of care to critical care unit | 11(32.4%) |
| Stepdown: | 6 (17.6%) |
| ICU: | 5 (14.7%) |
| Mechanical Ventilation | 4 (11.8%) |
| Mortality (of hospitalized/all PWH) | 1 (2.94%/0.97%) |

PWH-persons with HIV

## Discussion

This report describes the impact of COVID-19 infection on PWH at a single urban academic medical center in the US and identifies correlates of hospitalization. The study confirmed that hospitalization is related to host factors, such as older age and specific as well as multiple co-morbidities, and not to traditional metrics of HIV disease or immunosuppression. The median age of the cohort was similar to that in other studies and was lower than that of hospitalized HIV-uninfected patients with COVID-19.[3,8,10,11,12] The predominant co-morbidities for hospitalized patients—chronic lung and cardiovascular disease—were similar to those for patients in other COVID-19 cohorts. [8,13,14]

PWH are living longer with effective ART use, but are increasingly being diagnosed with new and multiple co-morbidities, particularly among communities of color.[15] While there is growing evidence that HIV-associated immunosuppression is not thought to be associated with COVID-19-related hospitalization or death, indirect measures of HIV and aging, as manifested in co-morbidities, still correlate with COVID-19 hospitalization. More recent reports from Moran et al.[16] from Grady Memorial Hospital in Atlanta, Georgia, found that co-morbidity burden was associated with hospitalization in a dose-dependent fashion when comparing hospitalized and nonhospitalized PWH with COVID-19. Sun et al. [17], in a National COVID Cohort Collaborative (N3C) study, looked at a large database of PWH with COVID-19 at US academic medical centers, compared with solid organ transplant patients, again noted that increased hospitalization and mechanical ventilation among PWH was related to the co-morbidity burden and not to specific demographics. However, the odds of hospitalization were observed to be higher for those with HIV than for the HIV-uninfected.

Despite well-maintained pre-COVID CD4 counts and suppressed HIVVL, HIV-associated immune dysfunction may affect the person’s ability to respond to the exuberant inflammatory reaction, resulting in clinical deterioration from COVID-19 [18]. Yendewa et al.[19] assessed a large health-care network of 44 facilities where PWH under less intensive ART and with much lower rates of viral suppression had higher odds of hospitalization, ICU admission, and mechanical ventilation, than patients without HIV infection. Co-morbidities were not reported, but the two groups of patients did have comparable 30-day mortality rates. Interestingly, among a subset of our hospitalized patients with CD4 counts performed after admission in this study, there was a significant drop in the absolute CD4 count consistent with nonspecific changes seen in acute illness.

The direct antiviral benefit of ART could be another explanation for less severe disease. A lower risk of hospitalization and severe disease was observed among other cohorts of patients on tenofovir disoproxil.[20,21] The current study, though limited by sample size showed no indication that receipt of ART had any effect on the clinical course or outcome. Differences in outcomes such as mortality may also have been affected by the variety of care standards used in each unique facility. Maintaining the current ART of patients as well as current standards of care, produces favorable outcomes for the overwhelming majority.

The study had several limitations. First, this was a retrospective review of PWH at a single urban institution, and this group may not be representative of the general global population of PWH. Second, hospitalized cases were identified through a health systems database with laboratory confirmation, leaving open the possibility that other PWH from the YNHH clinics were tested for SARS-CoV-2 or hospitalized elsewhere. Third, although reports show that COVID-19 disproportionately affects disadvantaged populations[22], the PWH cohort in the current study, consisting largely of minorities, and people over 50 years with multiple co-morbid conditions but with ready access to advanced treatments, achieved favorable outcomes overall. Finally, these data reflect the COVID-19 trends among PWH before the initiation of mass vaccination. As the pandemic continues to evolve and new variants emerge, these circumstances could alter the risk factors for hospitalization and severe disease.

## Conclusion

In this retrospective study, 103 PWH diagnosed with COVID-19 in an urban, resource rich academic center were identified. Compared with PWH managed as outpatients, those needing hospitalization were found to be older, with significantly more non-communicable co-morbidities. Mortality for the entire group was <1%. HIV-attributable factors were not associated with hospitalization for COVID-19.

## Data Availability

All data referred to in the manuscript is available for review.

## Acknowledgments

We are we grateful for the editing assistance of Mary-Ann Asico in the preparation of the report. Most importantly, we thank our patients.

